# The impact of the COVID-19 pandemic on end of life care in LTCFs in England: A qualitative study of LTCF staff experiences

**DOI:** 10.1101/2025.06.09.25328142

**Authors:** Natalie Cotterell, Danni Collingridge Moore

## Abstract

**Background:** The COVID-19 pandemic significantly increased the number of deaths within LTCFs globally. Restrictions around visitation and social distancing were common, however, research conducted during the pandemic demonstrates that these policies impacted the ways in which end of life care was delivered in LTCFs.

**Aim:** This paper aims to understand the experiences of LTCF staff in providing end of life care in the UK and explores the barriers and facilitators to doing so in the context of policies issued by the government at the time.

**Methods:** Data from semi-structured interviews conducted with 24 LTCF staff working across eight LTCFs in the north-west of England were analysed. Qualitative interviews were conducted with LTCF staff members, exploring their experiences of working in adult social care during the COVID-19 pandemic. Themes related to providing end of life care during this time were identified and analysed using thematic analysis.

**Findings:** Thematic analysis identified four key themes including: discrepancies in following COVID-19 UK government guidelines including visitation at end of life as an exception; the influence of staffing on delivering end of life care; utilising technology to substitute physical presence at end of life visits; and the emotional impact of delivering end of life care under COVID-19 restrictions.

**Conclusions:** The findings demonstrate the numerous challenges care staff experienced when delivering end of life care during the COVID-19 pandemic in terms of the practicalities of managing resident deaths, facilitating visitation and the associated impact on emotional wellbeing. Ensuring that all LTCF staff are trained to recognise end of life care, in the event of a future pandemic, will better equip LTCFs. In addition, it is paramount that the government provide consistent guidance on managing family contact at end of life, while taking into account the impact of implementing such guidance on the mental and emotional wellbeing of LTCF staff members.

## Introduction

The COVID-19 pandemic had a substantial impact on older adults living in long-term care facilities (LTCFs). During the course of the pandemic, the number of deaths occurring in LTCFs across the world significantly increased (1). In England, between March 2020 and January 2022, 43,256 death certificates of LTCF residents included COVID-19, accounting for 16.6% of all LTCF resident deaths (2). In addition to the LTCF deaths from COVID-19, residents were frequently deemed unsuitable for hospital admission due to pressure on bed occupancy, regardless of COVID-19 status, which, alongside limitations on the movement of residents, staff and health professionals between settings, contributed to end of life care increasingly taking place within LTCFs (3). Unsurprisingly, the COVID-19 pandemic had widespread implications for the delivery of end of life care in LTCFs, both for residents infected with COVID-19 and those who died from other causes.

The terms ‘palliative or ‘end of life care’ are often used interchangeably in Europe (4); in this paper, the term ‘end of life care’ will be used to refer to the care that individuals receive when they are likely to die within the next 12 months, with the aim of relieving suffering and improving quality of life for patients and families dealing with any type of life-threatening illness (5). Prior to the pandemic, the UK National End of Life Care Strategy recognised the need for high quality end-of-life care in LTCFs, however, delivering such care in these settings can be difficult (6). Barriers to implementing end of life care interventions include limited recognition of the need for end of life care, lack of support from management or wider stakeholders, and difficulties adopting end of life care into routine practice (7). These are additional challenges to the wider issues encountered in providing end of life care to older adults and those with dementia, both of which are common characteristics of the LTCF population.

The impact of COVID-19 on end of life care in LTCFs is only now, in retrospect, being fully understood. At this time, LTCFs were expected to follow the COVID-19 restrictions relating to infection prevention and control, social distancing, and visitation, further limiting LTCF staff time and resources to deliver end of life care (8). Despite the increased demand on staff time, there was relatively little guidance on delivering end of life care during pandemics in LTCFs globally (9). One online national survey of LTCFs in England highlighted variation in the provision of palliative and end of life care, with 18% of LTCFs reporting that they did not allow visitors for residents approaching end of life (10). Although technology was used to support remote communication at end of life, qualitative studies have found this to be time consuming and difficult for residents to engage with, especially those living with dementia (11, 12). Furthermore, a mixed methods systematic review demonstrated the negative impact of the pandemic on end of life care in LTCFs (13). On reviewing nine studies, the authors found that end of life care, especially advance care planning, had been disrupted during the COVID-19 pandemic in the UK, Ireland, Italy, Spain, Sweden, Peru, Mexico, and the United States. The authors identified four themes: the importance of advance care planning; increased responsibilities and expectations; emotional trauma and fear among staff; and professional pride and resilience, concluding that the pandemic reduced advance care planning, while increasing care staff responsibilities and exposure to death, which contributed to the heightened emotional trauma experienced by staff. These findings are consistent with wider research that has demonstrated the overall emotional toll that residents’ deaths during the pandemic had on LTCF staff (14, 15). Initiatives to provide online learning on end of life care during the pandemic resulted in increased awareness and understanding of advance care planning, increased confidence for advance care planning, and increased willingness to talk about advance care planning, however staff engagement was limited by a lack of time to complete the training (16).

The current paper draws upon data collected as part of a wider study which aimed to explore the implementation of COVID-19 restrictions in LTCFs in north-west England, examining the extent to which policies issued by the government were effective in their implementation. An analysis of data collected during semi-structured interviews with LTCF staff working during the pandemic on the impact of the COVID-19 pandemic specifically on end of life care is presented here. The analysis aims to understand the experiences of LTCF staff in providing end of life care and explore the barriers and facilitators to doing so in the context of wider government issued guidance in place at the time.

## Methods

### Research design

The findings presented in this paper are based on the secondary analysis of data from semi-structured, qualitative interviews with LTCF staff that were undertaken as part of a larger mixed-methods case study. The study aimed to explore the implementation of government issued policies in LTCFs during the COVID-19 pandemic. Further reporting of the methodological approach used has been reported elsewhere (17).

### Recruitment and sampling

Data collection took place in eight LTCFs in north-west England, within six local authorities. LTCFs were eligible to take part if they were registered with the Care Quality Commission (CQC) to provide residential or nursing care to older adults aged 65 and over and the facility was active during the COVID-19 pandemic. LTCFs were notified of the study between October 2023 and March 2024 through the Enabling Research in Care Homes Network (18).

In each facility, between two to four staff were identified by the facility manager to be approached with information packs about taking part in qualitative interviews. Inclusion criteria included being able to participate in the interview in English, being aged 18 years or older, willing to provide written informed consent and previous experience working in the facility during the COVID-19 pandemic. Written informed consent was provided by the facility manager and interview participants prior to data collection. Ethical approval was obtained from Lancaster University FHM Research Ethics Committee (reference: FHM-2023-3368-RECR-3).

### Qualitative, semi-structured interviews

Semi-structured interviews were conducted either online using Microsoft Teams or in person (in a private meeting space within the LTCF). Interviews ranged from 40 to 60 minutes. The topic guide used in the interviews focused on the experience of LTCF staff members in implementing the key policy recommendations issued by the UK government during the COVID-19 pandemic. All interviews were carried out at a convenient time during each staff members working shift. Each interviewee received a £25 gift voucher as a thank you gesture. All interview data were anonymised upon transcription and participants were given unique identification codes. In this paper, facility identifiers have been removed from quotations to ensure individuals cannot be identified from the data reported.

### Thematic analysis

All interviews were audio recorded and transcribed verbatim using Microsoft Teams and managed using Atlas.ti. software (19). Transcripts were checked for accuracy by NC and DCM. The data collected were analysed using thematic analysis (20). In the first step, data familiarisation, the transcripts were read and re-read by the research team to allow for familiarisation. Secondly, initial codes were generated, and thirdly emerging themes were identified in discussion with the research team and were revised iteratively. Fourthly, these themes were reviewed and finally the remaining themes were defined and named [28]. The COREQ checklist for the comprehensive reporting of qualitative studies was used to guide the reporting of this analysis (21).

## Results

### Sample characteristics

Twenty four LTCF staff members participated in semi-structured interviews. Twenty one participants were female, and twenty one participants identified as White British (of the remaining three, two identified as Indian or British Indian, and one identified as Portuguese). Twenty two reported English as their first language, and the average age was 46 years, ranging from 23 to 68 years. Participants had spent between 5 to 46 years working in direct patient care (average 21 years) and 5 to 46 years working in LTCFs (average 18 years). Participation was open to any staff role, providing the other inclusion criteria were met, and the sample included a range of managers, care assistants, housekeepers, and activity co-ordinators.

### Findings

All eight of the LTCFs included in the study had experience of providing end of life care both prior to and during the pandemic. One LTCF experienced no deaths from COVID-19 and provided end of life care for only one resident, who did not test positive for COVID-19, when the restrictions related to the pandemic had eased. Overall, LTCF staff had varying experiences and opinions on how and to what extent the pandemic had impacted end of life care. Most reported that the UK COVID-19 restrictions had a largely negative impact on end of life care, whilst others felt that it had minimal impact on the end of life care provided. In total, four core themes were conceptualised: discrepancies in following COVID-19 UK government guidelines (including visitation as an exception); the influence of staffing on delivering end of life care (including the recruitment of inexperienced staff; the role of wider health services; and the skill of recognising end of life); utilising technology to substitute physical presence at end of life visits (including digital contact not being the same as physical visits); and the emotional impact of delivering end of life care under COVID-19 restrictions (including trauma and helplessness). An overview of the core and sub-themes including descriptions can be found in Table 1.

**Table 1.** Overview of core themes including description of theme and sub-themes.

| Core theme | Description of theme | Sub-themes |
| --- | --- | --- |
| Discrepancies in following COVID-19 UK government guidance | This theme is centred around the inequities experienced both across and within facilities due to differing leadership approaches and infrastructure. | a. Visitation as an exception |
| The influence of staffing on delivering end of life care | This theme focuses on how staff shortages impacted care staff teams and the delivery of end of life care, in addition to highlighting the variable quality of relationships with wider services across case studies. | a. Recruitment of staff inexperienced with end of life<br>b. Role of wider health services<br>c. Recognising end of life |
| Utilising technology to substitute physical presence at end of life visits | This theme explores the role of technology in enabling communication with family at end of life visits. | a. Digital contact 'not the same' as physical visits |
| The emotional impact of delivering end of life care under COVID-19 restrictions | This theme examines the emotional impact of witnessing increased mortality on care staff, highlighting the helplessness and lack of formal support. | a. Trauma and helplessness |

### Discrepancies in adhering to COVID-19 government guidelines

#### Visitation as an exception

Most LTCFs reported allowing visitation at end of life, which at times was viewed by some participants as breaching the public guidance issued by the government. Some managers considered end of life to be an exceptional circumstance, claiming it was ‘different’:

*“We did take every precaution that we possibly could, but if somebody was dying that was different, that was, you know…”*

P014, F, Deputy care manager

The manager explained that they thought the risks of allowing visitation with Personal Protective Equipment (PPE) did not outweigh the benefits of visiting a loved one at the end of their life; thus, the facility bent the rules and allowed visitation at the end of life as an exception:

> *“Yeah, of course we did [allow visitation at the end of life]. I mean, you can’t…I don’t know how any sane person could say, oh, you’re not coming in in case you bring COVID to someone who’s already dying. You know, what difference is that going to make? Some of the things we did in our LTCF, we probably shouldn’t have done, but in hindsight now I think that a lot of other LTCFs wish they’d done it that way because, you know, you’d think of yourself in that situation and you know, what are the risks really?”*

> P014, F, Deputy care manager

Other staff members spoke of how the organisation of the LTCF building helped them to allow visitation as an exception. One member of staff remembered how they moved residents who were at the end of their life to ground floor rooms that were more easily accessible, so they could ‘bend the rules’ and allow visitation under the exceptional circumstance of death being anticipated:

> *“We have three rooms on the ground floor and they have sliding doors onto the garden, so we did bend the rules for those people, so they were allowed one at a time, I think it was, to visit.”*

> P022, F, Registered manager

Another LTCF provided end of life care for two residents during the pandemic, both of whom were already living in conveniently placed rooms: one on the top floor by a fire escape and one on the ground floor next to the fire door. This enabled the LTCF to allow visitation as an exception at end of life with relative ease:

> *“It was really quite funny because the two people who died were at the end of the building. One was on the top floor, so the family used to come up the fire escape and come in. They’d walk up and then they come in that door, and then the other lady was on the ground floor in the room below, and there was a fire door right next to her, so she’d just come. She’d ring the front doorbell, and then she’d go round to where she needed to. She used to go just straight in the room with her face mask and the gloves and stuff and then just literally go back out again. She didn’t… they didn’t mingle, we said if you need anything or you’re worried about your mum or whatever, just press the bell and somebody will come.”*

> P002, F, Registered manager

Other LTCFs followed the guidance more strictly and did not allow visitation even when a resident was viewed as nearing the end of their life. Staff reported finding it difficult seeing relatives watching their loved one die from outside the facility, i.e., through a window; one member of staff revealed that they breached the guidance and hugged a grieving relative, claiming standing there and not providing comfort was ‘not a human thing to do’:

> *“There’s one person that sticks in my head. It was a lady who looked, she was in a bedroom here, on end of life and her family couldn’t come in, and they were stood at the window with the priest watching her die. And we were in the room, trying to hold it together. We couldn’t. We couldn’t hold it together. You wanted to hug the relatives, and say sorry, but you can’t. I’m not gonna lie though, there was one woman who I was very close to, and I’m very close to the family and she died and I said I can’t stand here and watch you break down on your own, and I did hug her because I can’t watch somebody physically breakdown and just stand there, it’s not a human thing to do. You can give me guidance but I’m going to support that person.”*
>
> P013, M, Activity coordinator

In another LTCF, a senior carer spoke about finding the rapidly changing guidance difficult to deal with in terms of allowing visitation at end of life, with one relative unable to say goodbye to their husband, and another being allowed to visit their husband at end of life just a two weeks later:

> *“Yeah, we had this one guy, and he was dying, and they wouldn’t let his wife come and see him, so she had to stand at his bedroom window. And then there was, like, two weeks later, another guy was dying, but then the rules had changed, his wife was allowed in, and they were friends, so then she was really upset. You know like she couldn’t come in and see her husband, but her friend could two weeks later, yeah it was really hard.”*

> P007, F, Senior care assistant

In the same LTCF, the housekeeper described how a relative had to watch her husband die through a window ‘in the freezing cold’ and how ‘she should not have had to do that’, further demonstrating the conflict staff felt about the government guidance related to end of life:

> *“A lady who was outside watching her husband dying, taking his last breath, and she shouldn’t have had to do that. Not through a window in the freezing cold.”*
>
> P008, F, Housekeeper

### The influence of staffing on providing end of life care

#### The recruitment of staff inexperienced in end of life

All LTCFs who took part in the interviews reported that the pandemic caused staff shortages across their LTCFs. At the height of the pandemic, the government guidance advised care staff to isolate for up to two weeks if they developed symptoms of COVID-19 or if they or someone in their household tested positive for COVID-19 (8). In addition, managers reported losing members of staff after the government had announced that COVID-19 vaccinations were to be made mandatory for those working in LTCFs. This meant that some LTCFs relied on agency staff or rapidly recruited new starters, which had implications for end of life care within the facility.

Across numerous interviews, experiences of conflict between permanent LTCF staff and agency staff were reported. Several interviewees explained how agency staff did not know the building, residents, or routines well and how this impacted the quality of end of life care provided. Participant 001 described themself as ‘*a bit of a snob when it comes to end of life care’* as they carried out specific rituals when caring for individuals who had died. They lamented that the agency staff did not know the existing end of life routines within the LTCF, which made providing quality end of life care challenging:

> *“Like I said about the girl who had never seen a death. She’d never washed them. I’ve always been taught with my training as well, and what I like to do is I open the door to let the spirit out. I like to give them a wash, wear their favourite clothes and that’s how I’ve always been, and it’s like you’re doing one nearly every shift, and some of these people have never seen one before, because you come into here and you don’t necessarily die straight away… So we got a random agency, lovely people, didn’t know the routines you know, which was hard, but that’s what you’ve got to take, the stuff that you can.”*

> P001, F, CHAP team leader

Another care worker reported that the LTCF they worked in had rapidly recruited individuals who had no prior experience or training in providing social care – the majority of whom came from the hospitality sector as many employees were out of work due to the closure of hospitality establishments. Participant 009 reported that they felt those staff were more of a hindrance than a help as a significant amount of time was needed to show them what to do when workload was already overwhelming. In particular, end of life care was found to be a challenge for inexperienced staff who had not dealt with death before:

> “*We weren’t able to provide that* [end of life care] *in the best way because of shortage of staff. Another thing is the staff who started were quite new because they were working in a different job, for example, they were working in Asda […] ‘–’’But at that time, I was working like we were working with four staff for 35-36 residents, four staff and most of them (residents) were bed-bound. […] I had no time to teach them…but the company couldn’t find staff.”*

> P009, M, Unit manager

#### The role of wider health services

The relationship between LTCFs and wider health services varied greatly, especially in relation to the role of wider health services when delivering end of life care. Several LTCFs reported reduced support from wider services, forcing staff to take on more responsibilities around end of life. For example, staff reported having to increase their knowledge of end of life protocols and procedures, some of whom were asked to pronounce residents as dead with a physician or general practitioner present on the phone, at times when medical professionals avoided entering LTCFs:

> *“…even if you weren’t fully trained in how to pronounce someone dead, they’d ask you to do it on the phone. I did it on the phone to the doctor. Because they don’t want to come in, or if they did, they came in the spacesuit. […] We didn’t see many GPs (general practitioners) at all.”*

> P001, F, CHAP team leader

Several staff reported areas of potential bad practice from colleagues within the wider health service related to residents at end of life. One example was the use of blanket ‘Do Not Resuscitate (DNR)’ orders which were issued through the post to all residents in one LTCF without any prior discussion with the facility manager:

> *“We got loads of DNRs in the post from the doctors rolled up. Not individually or even named to that person […] and you opened it and there’s all these purple forms for loads of residents. Because that’s all the AMP’s* [Approved Medical Practitioner] *want to do as well. Just blanket caught everybody with DNR so they don’t have to do anything because we don’t matter.”*

> P013, M, Activity coordinator

The same interviewee recalled overhearing a conversation with a hospital doctor explaining to a resident that a DNR meant that they did not need to go into hospital if they tested positive for COVID-19:

> *“We got a lot of DNRs sent in the post. When there was… when people were explaining, and the doctors were ringing up… I’ll never forget this… they explained to a lady, who had dementia…well, she had memory loss, and he said to her, so we need to speak to you about do not resuscitate, and she said, what does that mean? He said that you don’t have to go to hospital, that’s what that means, and she agreed to that, to not being resuscitated so they lied, the doctors lied about what a DNR actually meant. They told her it just means you don’t go to hospital, if you get COVID. .”*

> P013, M, Activity coordinator.

A member of staff from another LTCF reported the restrictions on the resources LTCFs could have compared to wider health services meant that essentials such as oxygen were not available. They suggested that negative societal attitudes towards older people prevented residents from receiving appropriate healthcare, demonstrating the lack of support LTCFs received from wider services:

> *“I had to try and comfort people at the time who were dying. We couldn’t have oxygen. We were not allowed to. We were not allowed to have oxygen. They weren’t considered suitable to go to hospital because hospital was full, and we were told that they were old, and they had to stay here. […] What was the heartbreaking thing was one resident got refused hospital and oxygen. Nobody should be refused oxygen. Nobody.”*

> P006, F, Registered manager

#### Recognising end of life

Several LTCF staff discussed the speed and variability of decline of individuals dying with COVID-19, reportedly making it difficult to recognise when a resident was about to die. This was a challenge as staff needed to recognise when someone was ‘really at the end of life’ so that they could consider informing family members, allowing and organising visitation. When Participant 011 was asked whether many residents died alone, they spoke about the difficulty of ‘getting there on time’ when individuals were COVID-19 positive as the decline was often quick and there were minimal staff to facilitate visiting:

> “*We tried to get there, you know, we did try to get there. I mean, the last thing you want is for somebody to die on their own. That’s the very last thing you want. We do try to get there, but like I said, we were running on minimal staff, and I’ll be honest, I don’t know how we did it.”*

> P011, F, Deputy care manager

Participant 006 explained how COVID-19 more commonly infected those who were already nearing the end of their life, detailing how they could recognise that an individual was going to die from COVID-19:

> *“You could see the ones that were going to die because they just turned blue all of a sudden, they just turned blue. It was a sight to see, you know […] it (COVID-19) did like to go into people that were end of life. It did seem to know.”*

> P006, F, Registered manager

Others reported the importance of developing skills to recognise the presentation of the stages of end of life that older adults commonly experience. Participant 019 reported that staff needed to recognise when individuals were in the last days of dying in order to allow visitation, explaining how many of their residents could be seen as at the end of their life and so it was a skill to recognise when visitation was necessary:

> *“It was more right at the beginning where it was really strict* [with no visitation]*, then as the restrictions eased, people could come in for end of life. But it really was in last days of dying, because actually a lot of our people, you could see as end of life, but it was in that last days of dying that people could come in. I seem to think there were restrictions in numbers. I’m sure it was. Yeah, funny I can’t remember it now… and now, of course, we would do it very differently as a service.”*

> P019, F, Clinical team manager

### Utilising technology to substitute physical presence at end of life visits

#### Digital contact ‘not the same’ as physical visits

All LTCFs used digital technology as a substitute for physical visitation during lockdowns, though only some used technology to aid communication for those residents at end of life. Those who did not utilise technology instead facilitated face-to-face visitation when practically possible. In the LTCFs who did not offer visitation as an exception at end of life, digital technology was used to assist communication between the resident and their loved ones. Most LTCFs received dedicated iPads to assist with communication; however, in others where iPads were not purchased, staff were forced to use their personal phones.

Several staff, however, commented on how they felt it was inappropriate to substitute physical visitation with digital communication at end of life, particularly as residents at end of life often could not look at the screen, comprehend the nature of the communication, let alone understand how to use the technology effectively. Participant 010 felt communication through technology was ‘totally different’ to having a physical presence:

> *“How far the video call and virtual would support an end of life person rather than if somebody had been in and touched their hand is totally different to someone saying hi over the phone or a video call.”*

> P010, F, Clinical manager

Other staff members echoed this point; Participant 018 spoke of the difference between physical and online communication between end of life residents and their loved ones, highlighting that their LTCF ‘safely’ breached public guidance and prioritised face-to-face visiting for those at end of life:

> *“Obviously we tried to facilitate things like Zoom, but you know when you’re nonverbal and you know… there’s nothing like that contact really, so we really stretched the special circumstances kind of thing and used it to our advantage really, but also was kept safe in doing that, but making sure that people got that family time.”*

> P018, F, Head of operations

The presence of physical contact at end of life was highlighted as important by all interviewees. Participant 001 described the heartbreak they felt when assisting video calls with dying residents, reporting how they grew closer with the residents’ families through empathising with their situation:

> *“Not even ones that were dying were allowed to have any visitors, which was heartbreaking. It was just heartbreaking and watching them, like seeing their parents or grandparents dying on a phone like it’s I’m really sorry, but no it’s not gonna be the night, kind of thing. It’s do you want me to video call you? It’s how to reassure someone that’s dying on a video call and you not being able to be there and that’s where we got close with all the families.*”

> P001, F, CHAP team leader

### Emotional impact of delivering end of life care under COVID-19 restrictions

#### Trauma and helplessness

Staff who had provided end of life care during the pandemic or were present for a large number of deaths reported feeling traumatised by the experience, and there was a strong sense of helplessness amongst staff. Participant 012 echoed this, while Participant 006 reported that this sense of helplessness was also felt by the residents and relatives:

> “*It was tough because you’ve got so many and it’s like an impossible task because you just got so many just dropping in front of you.* […] *I felt helpless.”*

> P012, M, Senior care assistant

> *“They were helpless. I was helpless. The family, you know, the residents, those deaths were just horrendous, just horrendous.”*

> P006, F, Registered manager

Participant 001 described how traumatising they found some of the COVID-19 related deaths, mentioning that they had a younger member of staff who had started the job just weeks before the pandemic began and had no experience of seeing death:

> *“The amount of residents that we lost, there’s always one that’ll stick in my mind because it was traumatising, and that’s the girl I worked with who had never witnessed a death before. She’d never even done care before. Oh, it was awful. […] But yeah, it sounded like she* [the resident] *was gargling fluid like it was horrible. […] Oh I’ll never forget her, just how she was. It was just horrible, to think that’s what that virus did. It’s horrible, horrible.”*

> P001, F, CHAP team leader

One interviewee had received counselling after dealing with a significant number of deaths in a matter of weeks; others explained that they were still coming to terms with feeling helpless and not being able to prevent the deaths as Participant 013 echoed here:

> “*People’s lives were lost and there was nothing we could do about it. I still have nightmares to this day. There’s one that’s a recurrent one where I come into work, and all the residents are there that died. I see them all sat there, and they turn around and say ‘you let us die’, and I know I didn’t let them die, but that’s how it feels that we didn’t get the help. We didn’t get the help to help these residents and all we could do was just sit there with them while they died.”*

> P013, M, Activity coordinator

Overall, however, there was a lack of formal support for those who had provided end of life care during the pandemic, both in an emotional and practical sense. Participant 021 described how they were ‘left’, which further contributed to the challenges of providing appropriate end of life care:

> *“There was no support for us at all, there was nothing for any of the staff or certainly the people managing the situation. We were completely and utterly left on our own to it. […] It was horrible, yes. […] It made it even more difficult to give the best care, especially for people who were dying.”*

> P021, F, Care manager

## Discussion

The findings of this paper further expand the current knowledge base on the impact the COVID-19 pandemic had on providing end of life care in LTCFs, identifying four core themes that centre around: discrepancies following COVID-19 restrictions across LTCFs; the influence of staffing on the delivery of end of life care; the role of technology at end of life; and the emotional impact of delivering end of life care during the pandemic amongst care staff.

The themes identified support previous research demonstrating that service provision varied across LTCFs, subsequently impacting staff’s ability to provide end of life care (3). Staff shortages meant that LTCFs relied on agency staff and/or newly recruited, inexperienced individuals to deliver end of life care, which often contributed to the overwhelming workload of existing staff. In general, wider health services did not adequately facilitate appropriate end of life care within LTCFs, and bad practice was observed numerous times, for example, in the issuing of blanket DNRs, which had been highlighted as an area of concern early on in this pandemic (22). In addition, LTCFs reported widespread issues related to not receiving healthcare essentials such as oxygen and PPE, which prevented LTCFs from delivering appropriate end of life care. Furthermore, the lack of involvement from wider health services in LTCFs during the pandemic meant that staff responsibilities and roles at end of life increased. In particular, the importance of staff being able to quickly and confidently recognise when a resident was approaching end of life, subsequently allowing the facility to make decisions around visitation, was apparent. This was especially so for individuals who were approaching end of life and tested positive for COVID-19, as it was felt that these residents had variable symptoms and declined quickly compared to those without the virus.

LTCFs differed in the way they implemented the COVID-19 government guidelines; consistent with previous surveys, the majority of LTCFs allowed visitation as an exception at end of life, consequently breaching public guidance (10). The current research expands on this knowledge, demonstrating that this tended to happen in LTCFs with suitable building infrastructure which supported ‘safe visiting’, for example, where rooms were able to be accessed via fire escapes. Other LTCFs did not make any exceptions, meaning residents died alone, which staff reported as emotionally challenging and often had conflicted feelings about their role within this approach. Expanding upon this, staff further reported that relatives of residents found it difficult to see LTCFs implement different approaches to visitation at end of life, highlighting issues around equity.

The findings also demonstrate the importance of technology for end of life residents during the pandemic. Some LTCFs substituted all visits with digital technology, and although staff explained how technology was not the same as having a physical presence for dying residents, it was generally considered to be better than having no communication at all with loved ones before death. The adverse emotional impact of providing end of life care under COVID-19 restrictions was apparent across all staff members including long-term trauma due to experiencing feelings of helplessness. Supportive of previous research, staff were not offered or could not access adequate emotional support (23). Some staff mentioned that a lack of formal support, both emotional and practical, made it even more challenging to deliver appropriate end of life care. This heightened responsibility was particularly felt by managers who had emotionally vulnerable staff increasingly relying on them for emotional support. Previous research found that UK LTCF managers reported experiencing the highest levels of stress and anxiety during the pandemic compared to other European countries, increased responsibility and an overwhelming workload while dealing with a high number of resident deaths may be a potential contribution (24).

There are several implications of these findings, the majority of which in some form reflect changes that would benefit from being priority areas to address before the next pandemic. Firstly, the findings highlight the importance of having clear and consistent guidance on how LTCFs should operate during a pandemic, particularly addressing the delivery of end of life care. Much of the responsibility during the COVID-19 pandemic lay with the individual LTCF managers, meaning that the implementation of guidance varied significantly, unintentionally creating inequities amongst residents, staff, and relatives. Within this guidance, the potential emotional impact of implementing such guidance must be considered, emphasising the need for stakeholder involvement at all stages of policy development. In addition, any policy developed should be applicable in the context of end of life or have in built flexibility in its implementation when a resident is approaching end of life. These ‘caveats’ should be included within all aspects of protecting adult social care, including infection prevention and control measures, testing and visitation restrictions.

Secondly, there is a clear need for LTCF staff to be trained on end of life care, with a specific focus on identifying end of life in older adults. It is vital that LTCF staff can deliver appropriate end of life care; meaning that all staff should receive some form of training on how best to provide end of life care, including in the context of when resources such as staffing or access to wider health services are limited. Training could also include clear guidance on what the responsibilities of LTCF staff are, and where these responsibilities end, specifically in terms of the legalities of advance care planning. Staff time is often a barrier in such high pressure work environments; therefore, it is vital that individuals who work in LTCFs are consulted in the development of education resources and that any training is co-designed to increase the feasibility of implementing training within the LTCF (25).

Thirdly, it is important that LTCFs encourage residents to create informed advance care plans so that their wishes and goals of care, in the event of another pandemic, can be met. The importance of advance care planning has been highlighted in previous research and may help to avoid unethical and inappropriate blanket policies, such as issuing DNR notices to all residents, as discussed in this paper (26). There is also further potential for strategies and approaches that target and track advance care planning discussions for residents who may potentially be at a greater risk of contracting COVID-19, which have yet to be fully explored in England (27). The experiences of the COVID-19 pandemic also highlight the potential for discussing what to expect at end of life with residents and relatives, and how this could be handled by the LTCF, including if there were visitation restrictions in place, or if the resident deteriorated unexpectedly or at speed. Initiatives to support LTCF staff in communicating likely trajectories of decline at admission could support this (28).

Finally, there is an urgent need for emotional support for LTCF staff who continue to provide end of life care for residents. Long-term and unprocessed trauma was common amongst interviewees, with many stating that numerous colleagues had left the care sector completely either during or after the pandemic. The sector is characterised by a low-paid and often under-recognised workforce who deliver care to individuals with multiple and complex needs; yet the emotional demands of this work are too often forgotten (29). It is therefore recommended that care staff are offered appropriate emotional and bereavement support in order to process the COVID-19 pandemic, but also to build resilience and coping skills in the event of another pandemic (30).

### Strengths and limitations

This research is one of the first retrospective studies to be completed in LTCFs in England which considers the full timeline of COVID-19 restrictions implemented by the UK government, and its impact on the delivery of end of life care. In addition, data were collected in the context of interviews discussing wider policy recommendations, allowing the relative importance of how different policies during the pandemic to be directed by interview participants. There are, however, several limitations including: the authors were not able to interview individuals who resigned during the pandemic about their experiences of providing end of life care, meaning that the views of those who potentially left the long-term care sector are not reflected in the findings. Furthermore, all LTCFs that took part were most recently rated ‘good’ or ‘outstanding’ by the CQC, limiting the conclusions which can be drawn. The data also did not capture the level of staff knowledge or practices related to end of life care within each facility prior to the pandemic at a baseline, however the extent to which this devalues the experiences shared by staff members is limited.

## Conclusion

Overall, these findings have further explored how the COVID-19 pandemic adversely impacted the delivery of end of life care in LTCFs in England. LTCF staff were required to provide end of life care within the context of visitation restrictions, staff shortages and limited guidance. There are clear recommendations on how end of life could be managed better in future pandemics, however many of these require further training, investment, and support to be put in place now, in advance of its need. The emotional trauma staff endured during the pandemic while delivering end of life care is clear. In the future, it is imperative that research explores how best to support care workers, while amplifying the voices of adult social care workers is central to developing effective national responses to pandemics in the future.

## Data Availability

Data is available on reasonable request from the authors.

## Abbreviations

AMP: Approved Medical Practitioner
CHAP: Care Home Assistant Practitioner
CQC: Care Quality Commission
DNR: Do Not Resuscitate
GP: General Practitioner
LTCF: Long-term Care Facility
PPE: Personal Protective Equipment

## Ethics approval and consent to participate

Research Ethics Committee approval was obtained from the Lancaster University Faculty of Health and Medicine Research Ethics Committee (reference: FHM-2023-3368-RECR-3)

## Consent for publication

Written informed consent was collected from both the manager of the LTCFs recruited to the study and the LTCF staff member participating in the interview for anonymised publication of the data collected, included quotes.

## Data availability

Data is available from the authors upon reasonable request.

## Clinical trial number

N/A

## Authors’ contributions

DCM conceived the study, developed the study protocol, applied for ethical approval and contributed to the overall writing of the paper. NC coordinated recruitment conducted the interviews and prepared the manuscript. DCM and NC analysed the data.

## Acknowledgments

The authors would like to acknowledge and thank the Enabling Research in Care Homes network for supporting the recruitment for the study from which this data was collected. The authors would also like to thank the LTCFs and staff members who took part in this research for their valued contribution.

